# The occurrence and characteristics of severe pain in patients with axial spondyloarthritis

**DOI:** 10.1101/2022.01.31.22270160

**Authors:** Gareth T Jones, Ovidiu Rotariu, Linda E Dean, Alan G MacDonald, Gary J Macfarlane

## Abstract

**Objectives:** Pharmacological management of axial spondylarthritis (axSpA) seek to control inflammation. Even if successful, available evidence suggests that many patients continue to experience pain. The aim of the current study was to determine the prevalence and characteristics of severe pain, among persons with axSpA.

**Methods:** The Scotland Registry for Ankylosing Spondylitis (SIRAS) collected clinical and patient-reported data from adults seen in secondary care in Scotland with a clinical diagnosis of ankylosing spondylitis. Questionnaires asked about severe pain (high pain intensity; high pain interference; and extreme/unbearable pain), lifestyle, and various aspects of health. The relationship between severe pain and clinical/patient-reported factors was assessed using logistic regression.

**Results:** 929 participants had pain data available (73% male; median symptom duration 20yrs). High pain intensity and pain interference were more common (57% and 42%) than extreme/unbearable pain (11%). Prevalence did not differ with age, although women were less likely to report severe pain than men (Odds Ratios (ORs) 0.56-0.61) as were those with longer duration of education, and those from more affluent areas. The odds of severe pain increased with every 1 unit increase in BASFI (ORs 1.44-1.56). Strong associations were also seen with disease activity, spinal mobility, fatigue, poor sleep, and worse quality of life.

**Conclusion:** In axSpA, severe pain is common, with a clear socio-economic gradient and major impact on quality of life. Rheumatologists need to be aware of the large unmet need in terms of pain management in this patient group with around 1 in 9 patients reporting extreme/unbearable pain.

**KEY MESSAGES:** *What is already known about this subject?:*

- Axial spondyloarthritis is a painful inflammatory arthritis, although the epidemiology of specific pain phenotypes – and ‘severe’ pain in particular – is unknown.

*What does this study add?:*

- Severe pain (extreme/unbearable pain, pain of high intensity, or with a high level of interference) is commonly reported among people with axial spondyloarthritis.
- Three-quarters of individuals reporting severe pain still report severe pain two years later.
- Patients with severe pain are characterised by certain demographic characteristics, higher disease activity, poorer function, and a number of other clinical and lifestyle factors.

*How might this impact on clinical practice or future developments?:*

- Rheumatologists should be aware of the large unmet need in terms of pain management in this patient group. Specific pain management strategies should be considered to complement therapies aimed at reducing inflammation.

## INTRODUCTION

Axial Spondylarthritis (axSpA) is an inflammatory arthritis affecting predominantly the axial skeleton. The disease is characterised by pain, stiffness, and inflammation in the spine and sacroiliac joints, and peripheral joint disease and extra-articular features – uveitis, inflammatory bowel disease, psoriasis – are not uncommon. The origins of pain in axSpA are debated, with a proposed mixed model of both inflammatory and neuropathic pain present in some patients [1;2].

The effect of pain in axSpA is clear, with demonstrable negative impacts on quality of life, mood, and fatigue [3;4]. In a large systematic review and meta-analysis, we have shown that around 1 in 6 patients with axSpA also meet criteria for fibromyalgia [5]. Previous work, in a large UK-wide cohort of patients, has shown that fibromyalgia is associated with poorer disease activity and function, and occupational impairment [6]. Further, although there is no demonstrable effect on the response to biologic therapy, these patients start with worse disease, and therefore end with worse disease, despite showing some improvement [7].

Although pain is a predominant feature of the disease, it’s unclear whether specific pain phenotypes exist amongst axSpA patients, how common these are, their prognosis, and whether there are common characteristics that define them. Furthermore, although non-steroidal anti-inflammatory drugs (NSAIDs) and biologic DMARDs have been shown to be effective in reducing pain in axSpA [8], there is little data on the extent to which patients treated with these agents continue to report pain. Thus, the aim of the current study was to determine the prevalence, medication use, prognosis, and characteristics of severe pain, among persons with axSpA.

## PATIENTS AND METHODS

The Scotland Registry for Ankylosing Spondylitis (SIRAS) is a nationwide cohort of patients aged ≥16 years seen in secondary care in Scotland, with a clinical diagnosis of ankylosing spondylitis [9]. Patients were recruited from NHS hospitals between October 2010 and October 2013. Clinical data was extracted from patients’ medical records by trained research nurses and patients were sent postal questionnaires for self-completion. Follow-up questionnaires were sent at 1yr and 2yrs, although there was no further clinical data collection.

From questionnaire data participants were classified, in terms of presence/absence, with respect to three severe pain phenotypes:

1. High pain intensity – Any person scoring ≥50/100 on the Chronic Pain Grade [10] pain intensity sub-scale.
2. High pain interference – Any person with either:
  a. A score of ≥5/10 on the Chronic Pain Grade question: In the past 6 months, how much has pain interfered with your daily activities? And/or
  b. A response of ‘extremely’, to the SF12 Health Questionnaire [11] question: How much did pain interfere with your normal work?
3. Extreme/unbearable pain – Any person reporting ‘extreme pain or discomfort’ on the EQ-5D quality of life questionnaire [12].

The questionnaires also collected data on pain-related sleep disturbance (number of nights in the past four weeks); fatigue (Chalder Fatigue Scale [13], scored from 0 (low) to 11 (high)); and quality of life (Ankylosing Spondylitis Quality of Life index (ASQoL [14], scored from 0 (high) to 18 (low)). Socioeconomic factors, including education level, employment status, smoking status and alcohol intake were also collected, and participant postcodes were used to derive an estimate of deprivation (the Scottish Index of Multiple Deprivation [15]) based on area of residence.

Clinical data extracted from participants’ medical records included: disease duration, HLA-B27 status, presence of extra-spinal manifestations (psoriasis, uveitis, inflammatory bowel disease and peripheral joint involvement), medication use, and Bath Indices for disease activity (BASDAI [16]), physical function (BASFI [17]) and spinal mobility (BASMI [18]), all scored 0 (best) to 10 (worst).

Participants’ baseline characteristics were examined using simple descriptive statistics appropriate to data type: median (inter-quartile range (IQR)) for continuous/count data, and proportions (expressed as percentages) for categorical variables. Thereafter, the prognosis of severe pain was determined by the prevalence of participants in each pain phenotype who remained in that phenotype over the two-year follow-up period. Factors associated with severe pain (at baseline) were assessed with descriptive statistics and then characterised using logistic regression. Thus, results are expressed as odds ratios (ORs) with 95% confidence intervals. Independent variables were modelled as continuous variables if data allowed or were otherwise categorised for analysis.

The measurement of disease activity (BASDAI) includes two questions about pain. Therefore, the total BASDAI score was re-computed omitting these two questions and then statistically adjusted back to a 0-10 scale (low disease activity, to high). Here, this variable is referred to as BASDAI_adj_.

The SIRAS study was approved by the North of Scotland Research Ethics Service (09/S0802/7). The current analysis followed a pre-specified protocol. The protocol is available via Open Science Framework (https://osf.io/jsqx9) where it was deposited on 11-Mar-2020, prior to analysis commencing.

## RESULTS

929 participants had pain data available and were eligible for this analysis: 73% male, with a median age and symptom duration of 53 (IQR: 44-62) years and 20 (12-31) years, respectively. Baseline demographic and clinical characteristics of the study population is shown in Table 1.

**Table 1:**
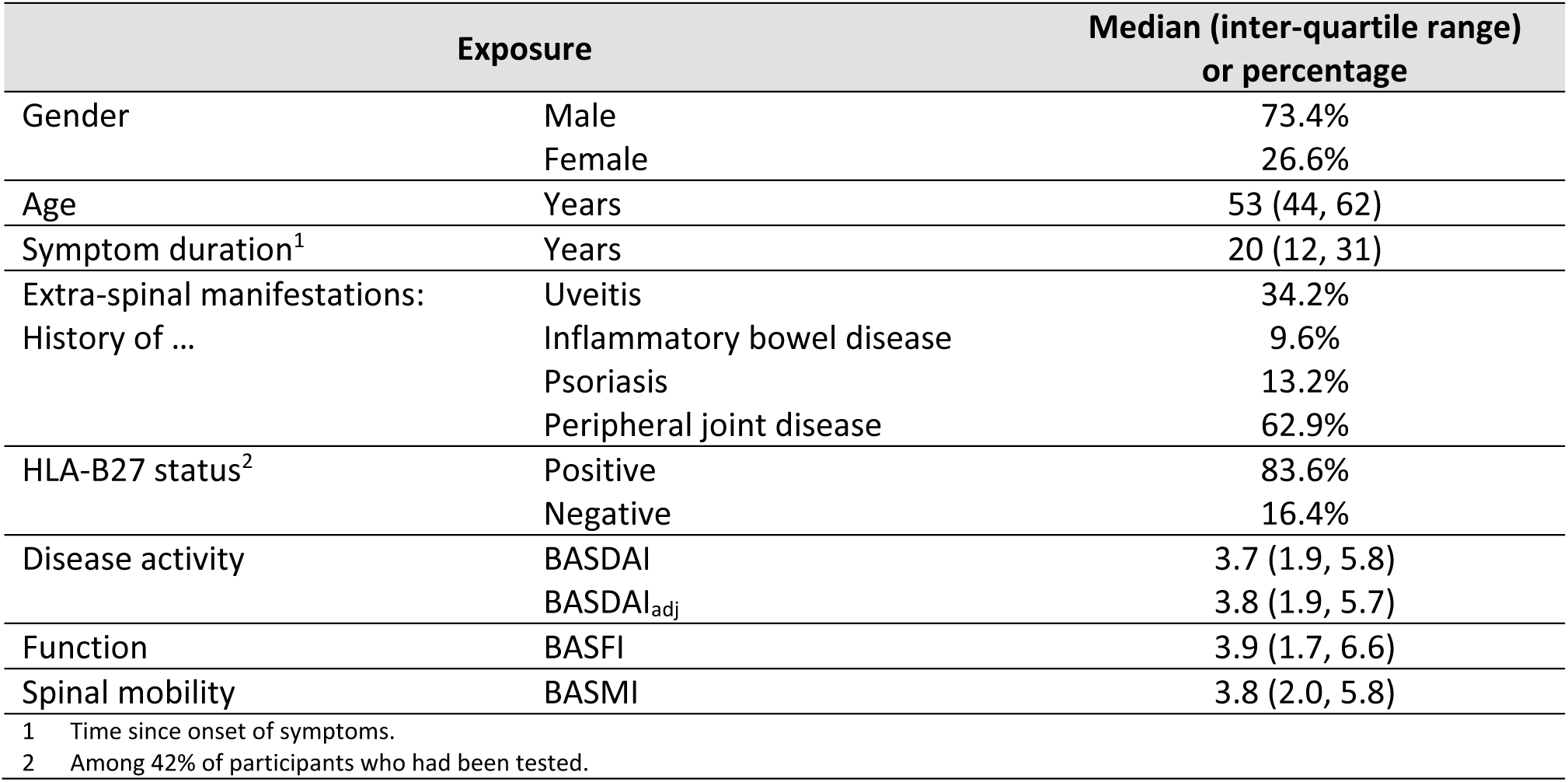
Participants’ baseline characteristics.

| Exposure |  | Median (inter-quartile range) or percentage |
| --- | --- | --- |
| Gender | Male | 73.4% |
|  | Female | 26.6% |
| Age | Years | 53 (44, 62) |
| Symptom duration<sup>1</sup> | Years | 20 (12, 31) |
| Extra-spinal manifestations: History of ... | Uveitis | 34.2% |
|  | Inflammatory bowel disease | 9.6% |
|  | Psoriasis | 13.2% |
|  | Peripheral joint disease | 62.9% |
| HLA-B27 status<sup>2</sup> | Positive | 83.6% |
|  | Negative | 16.4% |
| Disease activity | BASDAI | 3.7 (1.9, 5.8) |
|  | BASDAI<sub>adj</sub> | 3.8 (1.9, 5.7) |
| Function | BASFI | 3.9 (1.7, 6.6) |
| Spinal mobility | BASMI | 3.8 (2.0, 5.8) |

<sup>1</sup> Time since onset of symptoms. <sup>2</sup> Among 42% of participants who had been tested.

In total, 556 individuals (61%) reported severe pain satisfying at least one of the three phenotypes. Pain of high intensity (57%) and with a high level of interference (42%) were common, and the overlap between these phenotypes was great (Figure 1). Extreme/unbearable pain, although less prevalent, was still reported by a sizeable minority (11%) although was seldom reported in isolation. 95 participants (10% of the total population) met all three criteria.

**Figure 1:**
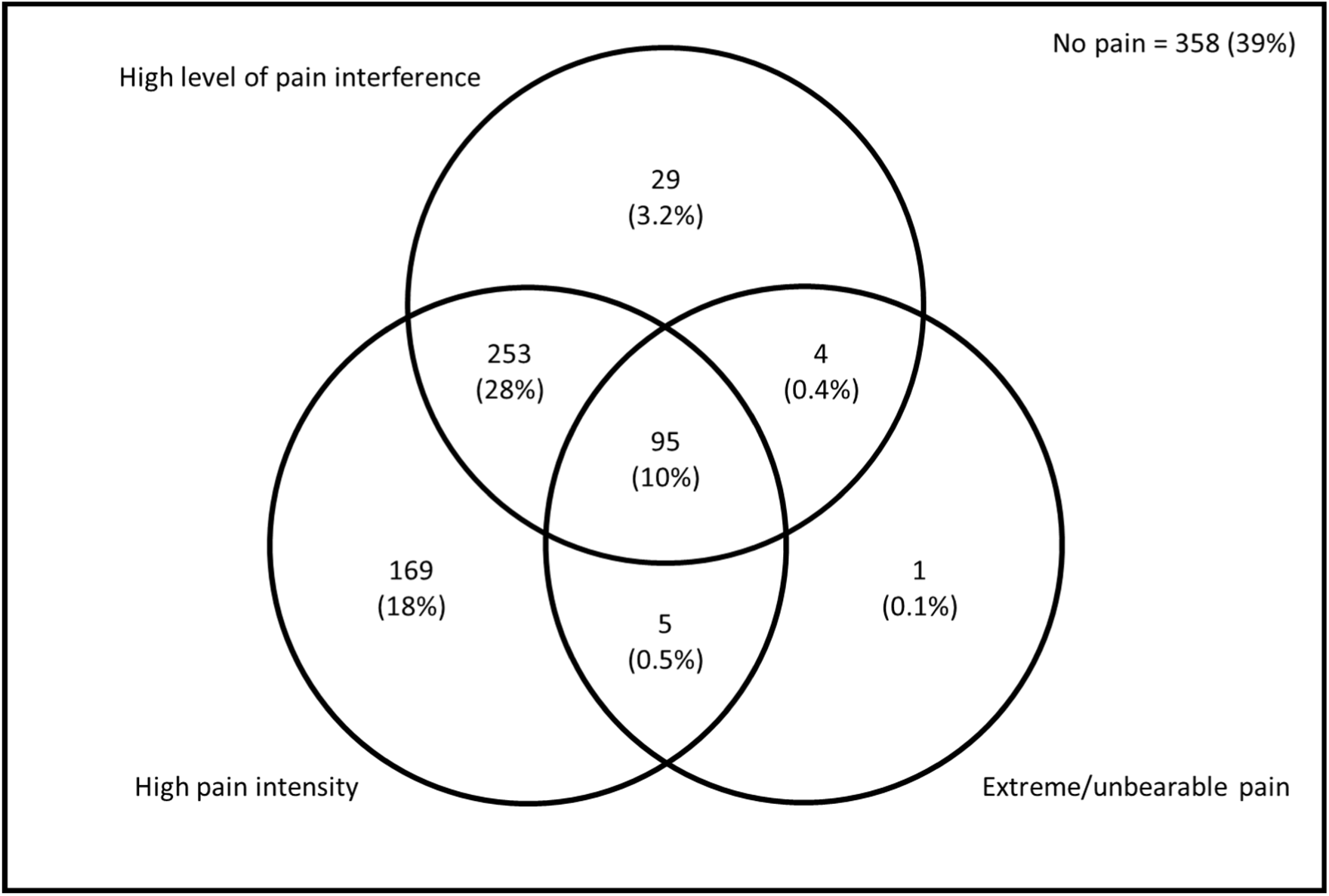
Overlap between severe pain phenotypes.

Table 2 describes participants’ medication use, stratified by pain phenotype. Medication use was common: 89.6% for the entire cohort, and slightly higher among those with severe pain. Participants with extreme/unbearable pain or high pain intensity were at significantly increased odds of being prescribed biologic therapy compared to patients not meeting these pain phenotypes (OR: 1.83; 95%CI: 1.17-2.88, and OR: 1.67; 95%CI: 1.20-2.33 respectively). A similar effect was observed among those with high pain interference, although the effect was more modest and not statistically significant (OR: 1.33; 95%CI: 0.97-1.84). Among participants taking any medication reporting severe pain was associated with a 21% to 49% increase in the odds of receiving combination therapy (extreme/unbearable pain (OR: 1.21; 95%CI: 0.78-1.88); high pain intensity (OR: 1.32; 95%CI: 1.003-1.75) and high pain interference (OR: 1.49; 95%CI: 1.12-1.97)).

**Table 2:** Medication use, stratified by severe pain phenotype.

| Exposure | High pain intensity |  | High pain interference |  | Extreme/unbearable pain |  |
| --- | --- | --- | --- | --- | --- | --- |
|  | N | (%) | N | (%) | N | (%) |
| Total | 522 |  | 381 |  | 105 |  |
| No medication | 43 | (8.2%) | 27 | (7.1%) | 10 | (9.5%) |
| **Monotherapy** |  |  |  |  |  |  |
| NSAID | 105 | (20.1%) | 73 | (19.2%) | 17 | (16.2%) |
| csDMARD | 13 | (2.5%) | 9 | (2.4%) | 2 | (1.9%) |
| Biologic agents<sup>1</sup> | 46 | (8.8%) | 32 | (8.4%) | 13 | (12.4%) |
| Other<sup>2</sup> | 23 | (4.4%) | 15 | (3.9%) | 4 | (3.8%) |
| **Combination therapy** |  |  |  |  |  |  |
| NSAID + csDMARD | 31 | (5.9%) | 27 | (7.1%) | 3 | (2.9%) |
| NSAID + Biologic | 53 | (10.2%) | 37 | (9.7%) | 11 | (10.5%) |
| csDMARD + Biologic | 7 | (1.3%) | 5 | (1.3%) | 1 | (1.0%) |
| NSAID + csDMARD + Biologic | 20 | (3.8%) | 15 | (3.9%) | 7 | (6.7%) |
| All other combinations<sup>3</sup> | 181 | (34.7%) | 141 | (37.0%) | 37 | (35.2%) |

Abbreviations. NSAID: Non-Steroidal Anti-Inflammatory Drugs. csDMARD: Conventional synthetic Disease Modifying Anti-Rheumatic Drugs. <sup>1</sup> Biologic therapies consisted solely of adalimumab (Humira), etanercept (Enbrel), and infliximab (Remicade). <sup>2</sup> Includes steroids. (Only 3 patients were taking steroid monotherapy.) <sup>3</sup> All other combination therapies, including combination with steroids.

Follow-up data was available for 487 (52%) of participants at 1yr, 301 (62%) of whom also provided data at 2yrs. Baseline pain intensity was highly indicative of pain intensity over the duration of follow-up with ≥74% of participants remaining in the same categories at 12 and 24-month follow-up. Similar trajectories were observed for high pain interference, and the presence of extreme/unbearable pain (see Supplementary Figures 1-3).

Severe pain did not differ with age. However, across pain phenotypes women were 40-50% less likely to report severe pain than men (OR range: 0.56 to 0.61) (see Table 3 and Supplementary Tables 1-3). Participants with more years of education, and those living in more affluent areas, also had lower odds of severe pain. Severe pain was also strongly associated with employment status. Compared to those in full-time employment, the odds of high pain intensity were six times higher among participants who because of ill-health had retired early or were unemployed (OR: 6.12; 95%CI: 4.05-9.24). Similar effects were observed with extreme/unbearable pain (7.98; 95%CI: 4.55-14.0) and high pain interference (9.95; 95%CI: 6.74-14.7).

**Table 3:** Association between baseline characteristics and severe pain phenotypes.

| Exposure |  | High pain intensity |  | High pain interference |  | Extreme / unbearable pain |  |
| --- | --- | --- | --- | --- | --- | --- | --- |
|  |  | OR<sup>1</sup> | (95%CI) | OR | (95%CI) | OR | (95%CI) |
| **Demographic and lifestyle factors** |  |  |  |  |  |  |  |
| Age | <40 years | 1.00 |  | 1.00 |  | 1.00 |  |
|  | ≥40 to <60 years | 1.08 | (0.74-1.59) | 1.06 | (0.72-1.55) | 0.95 | (0.55-1.65) |
|  | ≥60 years | 1.23 | (0.81-1.86) | 1.22 | (0.81-1.84) | 0.58 | (0.30-1.10) |
| Gender | Male | 1.00 |  | 1.00 |  | 1.00 |  |
|  | Female | 0.56 | (0.41-0.76) | 0.61 | (0.46,0.82) | 0.59 | (0.38,0.90) |
| Education | Secondary school | 1.00 |  | 1.00 |  | 1.00 |  |
|  | Apprenticeship | 0.71 | (0.46-1.10) | 0.65 | (0.42-0.99) | 0.64 | (0.34-1.21) |
|  | College<sup>1</sup> | 0.67 | (0.48-0.94) | 0.63 | (0.45-0.87) | 0.41 | (0.25-0.69) |
|  | University degree | 0.46 | (0.31-0.69) | 0.34 | (0.22-0.52) | 0.20 | (0.09-0.48) |
|  | Further degree | 0.27 | (0.15-0.49) | 0.32 | (0.17-0.61) | 0.25 | (0.08-0.83) |
| Employment status | Working full-time | 1.00 |  | 1.00 |  | 1.00 |  |
|  | Working part-time | 1.39 | (0.89-2.18) | 2.10 | (1.32-3.35) | 3.39 | (1.60-7.20) |
|  | Retired | 1.34 | (0.91-1.95) | 1.63 | (1.08-2.45) | 1.88 | (0.88-4.01) |
|  | Ill-health<sup>1</sup> | 6.12 | (4.05-9.24) | 9.95 | (6.74-14.7) | 7.98 | (4.55-14.0) |
|  | Other<sup>1</sup> | 3.47 | (1.70-7.09) | 3.88 | (2.04-7.38) | 2.17 | (0.70-6.72) |
| Deprivation (quintiles) | 1 (most deprived) | 1.00 |  | 1.00 |  | 1.00 |  |
|  | 2 | 0.56 | (0.31-0.99) | 0.59 | (0.35-0.99) | 0.98 | (0.50-1.91) |
|  | 3 | 0.33 | (0.19-0.56) | 0.36 | (0.22-0.58) | 0.47 | (0.24-0.94) |
|  | 4 | 0.34 | (0.20-0.58) | 0.28 | (0.17-0.46) | 0.63 | (0.33-1.20) |
|  | 5 (least deprived) | 0.27 | (0.15-0.46) | 0.22 | (0.13-0.36) | 0.35 | (0.17-0.75) |
| Smoking status | Never | 1.00 |  | 1.00 |  | 1.00 |  |
|  | Ex | 1.26 | (0.94-1.70) | 1.37 | (1.01-1.85) | 1.26 | (0.76-2.11) |
|  | Current | 1.99 | (1.38-2.85) | 2.44 | (1.72-3.48) | 2.77 | (1.67-4.62) |
| Alcohol consumption | Never | 1.70 | (1.06-2.71) | 2.19 | (1.40-3.44) | 3.16 | (1.75-5.71) |
|  | Ex | 2.25 | (1.54-3.30) | 2.96 | (2.06-4.23) | 3.61 | (2.25-5.81) |
|  | Current | 1.00 |  | 1.00 |  | 1.00 |  |
| **Clinical and patient-reported characteristics** |  |  |  |  |  |  |  |
| Symptom duration | <10 years | 1.00 |  | 1.00 |  | 1.00 |  |
|  | 10 to <20 years | 1.38 | (0.87-2.20) | 1.20 | (0.75-1.93) | 1.92 | (0.78, 4.75) |
|  | 20 to <30 years | 1.68 | (1.03-2.73) | 1.49 | (0.92-2.43) | 2.04 | (0.81, 5.14) |
|  | ≥30 years | 1.46 | (0.91-2.35) | 1.63 | (1.01-2.63) | 2.19 | (0.89, 5.40) |
| History of extra-spinal manifestations | Uveitis – No | 1.00 |  | 1.00 |  | 1.00 |  |
|  | Uveitis – Yes | 0.96 | (0.72-1.26) | 0.97 | (0.73-1.28) | 0.73 | (0.47, 1.15) |
|  | IBD<sup>1</sup> – No | 1.00 |  | 1.00 |  | 1.00 |  |
|  | IBD – Yes | 0.99 | (0.64,1.55) | 1.34 | (0.87-2.08) | 1.13 | (0.58, 2.21) |
|  | Psoriasis – No | 1.00 |  | 1.00 |  | 1.00 |  |
|  | Psoriasis – Yes | 0.88 | (0.60-1.30) | 0.90 | (0.61-1.34) | 0.50 | (0.24, 1.05) |
|  | PJD<sup>1</sup> – No | 1.00 |  | 1.00 |  | 1.00 |  |
|  | PJD – Yes | 1.60 | (1.22-2.10) | 1.53 | (1.16-2.01) | 1.29 | (0.83, 1.97) |
| HLA-B27 status | Positive | 1.00 |  | 1.00 |  | 1.00 |  |
|  | Negative | 1.59 | (0.90-2.81) | 1.68 | (0.98-2.88) | 2.80 | (1.38, 5.67) |
| Disease activity (BASDAI<sub>adj</sub>) | 0 to 3 (low) | 1.00 |  | 1.00 |  | 1.00 |  |
|  | 3 to <6 | 4.20 | (2.51-6.93) | 4.05 | (2.35-6.96) | 1.80 | (0.64, 5.11) |
|  | 6 to 8 (high) | 13.6 | (6.38-30.0) | 12.8 | (6.50-25.2) | 14.7 | (5.73, 37.5) |
| Function (BASFI) | Per unit | 1.48 | (1.38-1.59) | 1.56 | (1.45-1.68) | 1.44 | (1.30, 1.60) |
| Spinal mobility (BASMI) | 0 to 3 (good) | 1.00 |  | 1.00 |  | 1.00 |  |
|  | 3 to <6 | 1.51 | (1.05-2.18) | 1.80 | (1.23-2.63) | 1.88 | (1.03, 3.44) |
|  | 6 to 10 (poor) | 2.83 | (1.80-4.44) | 2.85 | (1.85-4.40) | 1.78 | (0.90, 3.52) |
| Fatigue (Chalder Fatigue Scale) | 0 to 10 (low) | 1.00 |  | 1.00 |  | 1.00 | <sup>2</sup> |
|  | 10 to 19 | 1.95 | (1.07-3.54) | 2.58 | (1.24-5.40) | 3.91 | (0.53, 28.8) |
|  | 20 to 33 (high) | 13.8 | (6.25-30.4) | 19.3 | (8.33-44.6) | 34.6 | (4.65, 258) |
| Pain-related sleep disturbance: number of nights in past 4 weeks | 0 | 1.00 |  | 1.00 |  | 1.00 |  |
|  | 1 to 3 | 2.57 | (1.62-4.09) | 1.11 | (0.63-1.96) | – | <sup>3</sup> |
|  | 4 to 7 | 6.44 | (4.16-9.98) | 3.70 | (2.37-5.78) | 5.01 | (1.05, 23.8) |
|  | >7 to 14 | 12.8 | (7.37-22.3) | 8.53 | (5.19-14.0) | 33.9 | (7.73, 148) |
|  | >14 | 19.5 | (12.4-30.7) | 10.8 | (7.17-16.1) | 68.0 | (16.5, 281) |
| Quality of life (EQ-5D) | ≥0.75 to 1.0 (high) | 1.00 |  | 1.00 |  | – | <sup>4</sup> |
|  | 0.5 to <0.75 | 8.06 | (5.76-11.2) | 9.77 | (6.30-15.1) | – |  |
|  | <0.5 (low) | 61.9 | (30.1-127) | 102 | (53.2-194) | – |  |
| Mental health (SF12 MCS<sup>1</sup>) |  | 0.93 | (0.91, 0.94) | 0.92 | (0.90-0.93) | 0.90 | (0.88-0.92) |

Raw data underpinning this table is shown in Supplementary Tables 1-3.
Abbreviations: OR = Odds ratio. College = Further education college. Ill-health = Retired early or unemployed due to ill-health. Other = In unpaid work, unemployed and seeking work, or student. IBD = Inflammatory bowel disease. PJD = Peripheral joint disease. MCS = Mental Component Scale.
2 Odds ratios calculated by adding 1 unit to each cell, to avoid singularities.
3 Odds ratio cannot be computed, therefore exposure category collapsed with 4-7 nights.
4 Odds ratios cannot be computed due to distribution of data.

Current smokers were more likely to report severe pain (OR range: 1.99 to 2.77) and compared to participants who reported that they drank alcohol, ex-drinkers (range of ORs: 2.25 to 3.61) and never drinkers (range of ORs: 1.70 to 3.16) experienced higher odds of severe pain.

Among the 42% whom had been tested, participants who were HLA-B27 negative were significantly more likely to report extreme/unbearable pain (OR: 2.80; 95%CI: 1.38-5.67). There was some evidence that participants with a history of peripheral joint involvement had higher odds of severe pain compared to other individuals (range of ORs: 1.29 to 1.60). However, there were no consistent or significant associations across the other extra-spinal manifestations: uveitis, inflammatory bowel disease, or psoriasis. Participants with high disease activity (BASDAI_adj_) were significantly more likely to report severe pain (OR range: 12.8 to 14.7). Severe pain was also strongly associated with poorer function, fatigue, sleep disturbance, poor mental health, and poor quality of life (see Table 3 and Supplementary Tables 1-3).

## DISCUSSION

We have shown that the prevalence of severe pain – as defined by high pain intensity, high pain interference or extreme/unbearable pain – is common among persons with axSpA. In the current study, 57% of participants reported pain of high intensity, 42% reported pain with a high level of interference, and 11% reported extreme/unbearable pain. Also, we have demonstrated that this pain is persistent, with three-quarters still reporting such pain up to two years later. It is associated with a number of demographic and clinical factors, and several adverse outcomes such as fatigue, low quality of life and poor work outcomes.

There are several methodological issues to consider. Firstly, the potential for selection bias. Patients were included from secondary care rheumatology departments in 13 out of 14 regional health boards in Scotland; we did not receive permission from one small island health board, and data was incomplete from one other health board. Otherwise, we believe that the invited participants consisted of all patients with a clinical diagnosis of ankylosing spondylitis seen in secondary care in Scotland in the two years prior to data collection and is therefore representative of patient population in routine rheumatology practice. We have previously shown that only one-third of patients are managed in rheumatology [19]. These patients are younger, and more likely to have a history of uveitis, inflammatory bowel disease and psoriasis – although neither age nor these extra-articular features were associated with severe pain in the current analysis. However, while this previous analysis showed that patients still managed in secondary care were more likely to be younger age at diagnosis (35 versus 38yrs), this difference is small and unlikely to have introduced any major bias in the current study. Although clinical information was available from 1886 SIRAS participants (76% male; median age and symptom duration 51yrs and 19yrs), pain phenotype data was only available from 929 questionnaire respondents (49%). However, in terms of basic characteristics these respondents were similar to the overall clinical population: 73% male, with a median age and symptom duration of 53yrs and 20yrs, respectively. Although not conclusive, this would suggest that selection bias into the study is not a major concern.

We have shown a strong association between severe pain and high disease activity: participants with a BASDAI_adj_ score ≥6 experienced more than a ten-fold increase in the odds of severe pain (OR range: 12.8 (6.5-25.2) for high pain interference, to 14.7 (5.73-37.5) for extreme/unbearable pain). Ordinarily, high levels of pain would be expected to influence BASDAI score because it contains questions about pain in the neck, back or hip (question 2) and joints other than neck, back or hip (question 3). However, in the current study these two questions were eliminated and the computation of the BASDAI_adj_ is based on the four remaining non-pain items: fatigue/tiredness, tenderness/discomfort, and morning stiffness severity and duration. BASDAI has been shown to have high internal consistency [20]. The high Cronbach’s alpha (0.87) doesn’t necessarily mean that the instrument is unidimensional, but it does suggest that the individual questions are closely related. It is possible therefore, that even after omitting the questions on pain, the BASDAI_adj_ is still driven, in part, by pain status.

It is important to remember that pain and disease activity are not objectively measured constructs. Rather, data on both are gathered by self-report and it may be that pain influences reporting behaviour, even if it does not influence disease activity directly. It would be informative to have had an objective measures of disease status (such as c-reactive protein), but this information was not available in the current dataset.

Another limitation is the absence of data on fibromyalgia. It may be that those with more severe pain are more likely to have fibromyalgia – a comorbidity previously shown to be common in axSpA. Estimates vary depending on the classification criteria employed – of both fibromyalgia and axSpA – but we demonstrated, in a recent systematic review and meta-analysis, a pooled prevalence of 16.4% [5]. Data from a UK-wide cohort has shown that patients who meet criteria for fibromyalgia report higher disease activity and poorer quality of life than patients without [7]. At the time of data collection, American College of Rheumatology ‘research’ criteria for fibromyalgia were not available, and fibromyalgia presence/absence was seldomly recorded in participants’ medical notes.

The SIRAS study is a pragmatic cohort of patients in routine clinical practice. To be eligible for inclusion patients were required to have a clinical diagnosis of ankylosing spondylitis but were not required to meet any disease classification criteria such as modified New York [22], or ASAS criteria [23]. Future research may wish to examine differences (if they exist) between different sub-groups of axSpA.

Future investigations should also attempt to ascertain the site(s) of pain. In the current analysis we were only able to consider only pain severity, and not pain location. A separate analysis (not shown) examined chronic widespread pain, as per the Manchester definition – pain in two areas in contralateral body quadrants, as well as the axial skeleton [21]. We found similar associations that were observed in the current analysis – i.e. strong associations between chronic widespread pain and demographic characteristics (gender, education, employment status and deprivation), with lifestyle (smoking and alcohol consumption) and with various clinical (disease activity, function) and patient-reported (fatigue, quality of life) characteristics. This would suggest that the relationships we present here are not restricted to pain associated with axial disease, but with pain more generally. Knowing the site of pain may also help inform whether the pain is likely to be of inflammatory origin.

We have not produced multivariable models. It was not our intention to produce clinical prediction models that best predict the different severe pain phenotypes with a parsimonious set of exposure variables. Instead, we were seeking to identify groups of factors that are associated with severe pain that might be informative in the clinic in terms of directing resources. Neither are we claiming that the associations reported here are causal. One needs to be wary about reverse causation when interpreting these findings. For example, we report strong associations between severe pain and employment status, in particular with unemployment or early retirement due to ill-health. These are likely to be consequences of severe pain rather than causes of it. Similarity the causal nature of the relationship between pain and fatigue is unclear – and indeed it is likely, to a certain extent, to be bidirectional.

The relationship between high pain intensity and poor quality of life is not surprising. However, the strength of the association is remarkable: participants with high intensity pain experienced more than a 60-fold increase in the odds of poor quality of life, and the association with high pain interference was even greater. For extreme/unbearable pain the ORs were not computable due to the distribution of the data: *all* participants with extreme/unbearable pain had very low scores on the EQ-5D quality of life instrument. This is a function of the fact that the definition of extreme/unbearable pain was based on the EQ-5D pain question. However, it is of note that nearly three-quarters of the 105 participants with extreme/unbearable pain had an EQ-5D score below zero, equivalent to a health state “worse than death”. This emphasises the importance of appropriate pain management strategies in this patient group.

The main message of this manuscript is one of unmet need. The SIRAS study is somewhat historic – it comprises patients with long-standing disease (median symptom duration 20yrs) recruited between 2010-2013. Many, therefore, will have been diagnosed and initially treated in the pre-biologic era. With the advent of biologics and other targeted therapies it may be argued that, today, patients are better managed – and this is undoubtedly true, at least in part. Nonetheless, given the young age of onset of axSpA, and the fact that not everyone responds to biologics, there will be many patients growing old with severe pain which is probably not adequately treated, either in primary or secondary care. We have shown that patients with severe pain are more likely to receive biologic therapy. The challenge for rheumatology is how best to manage those patients whose pain won’t respond to biologic therapy.

In conclusion, in this national cohort of patients with axSpA, unselected for disease activity or treatment escalation, we have shown that sizeable proportion of patients report severe pain. Even ‘extreme/unbearable pain’, thought to be a relatively rare pain phenotype, is reported by around 1 in 9 patients. Rheumatologists should be aware of the large unmet need in terms of pain management in this patient group and be mindful of the clear social gradient that suggests that there is an excess of severe pain in lower socioeconomic groups.

## Data Availability

The data underlying this article will be shared on reasonable request to the corresponding author.

## ACKNOWLEDGEMENTS / CONTRIBUTIONS / DATA AVAILABILITY

## Acknowledgements

We thank all the clinicians and research nurses who facilitated recruitment and data collection to the SIRAS study. We are indebted to the SIRAS steering committee, especially Professor Roger Sturrock (chair) and Dr David Marshall (vice-chair). We also thank staff from the SIRAS coordinating centre, in particular: Elizabeth Ferguson-Jones, Giles O’Donovan, Nabi Moaven-Hashemi and Flora Joyce. For this analysis, we also acknowledge the contribution of Harriet Dickinson (GlaxoSmithKline) in protocol development.

## Author contributions

GTJ, AGM and GJM were involved in the design and oversight of the SIRAS study. GTJ, OR, LED and GJM designed the current analysis. Analysis was undertaken by OR, and overseen by GTJ, LED and GJM. GTJ produced the first draft of the manuscript, and all authors revised it critically for intellectual content. All authors approved the final manuscript for submission.

## Protocol

The analysis followed a pre-specified protocol. This is available via the Open Science Framework: https://osf.io/jsqx9.

## Data availability

The data underlying this article will be shared on reasonable request to the corresponding author.

## SUPPLEMENTARY DATA (ONLINE)

**Supplementary Figure 1:**
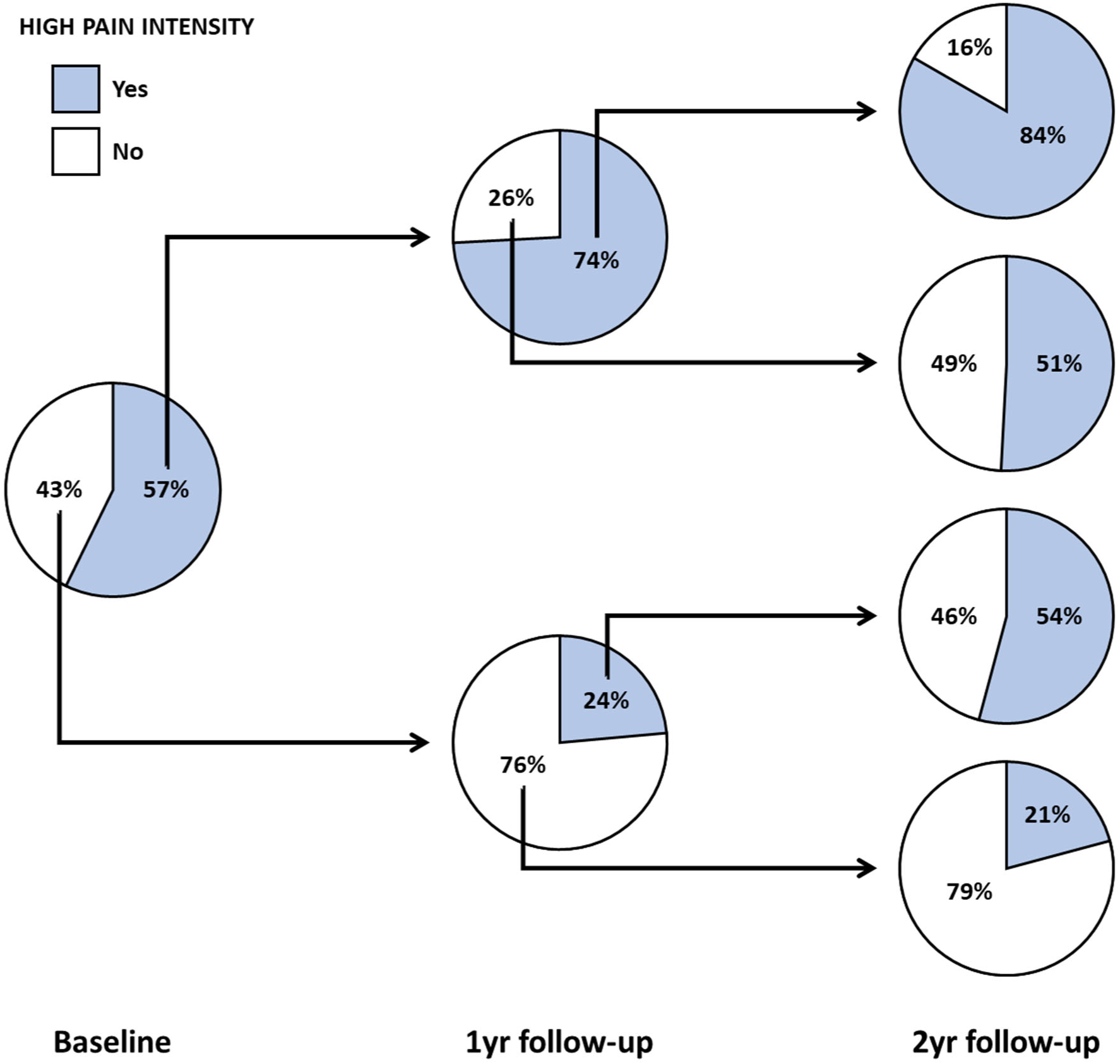
Longitudinal trajectory of high pain intensity.

**Supplementary Figure 2:**
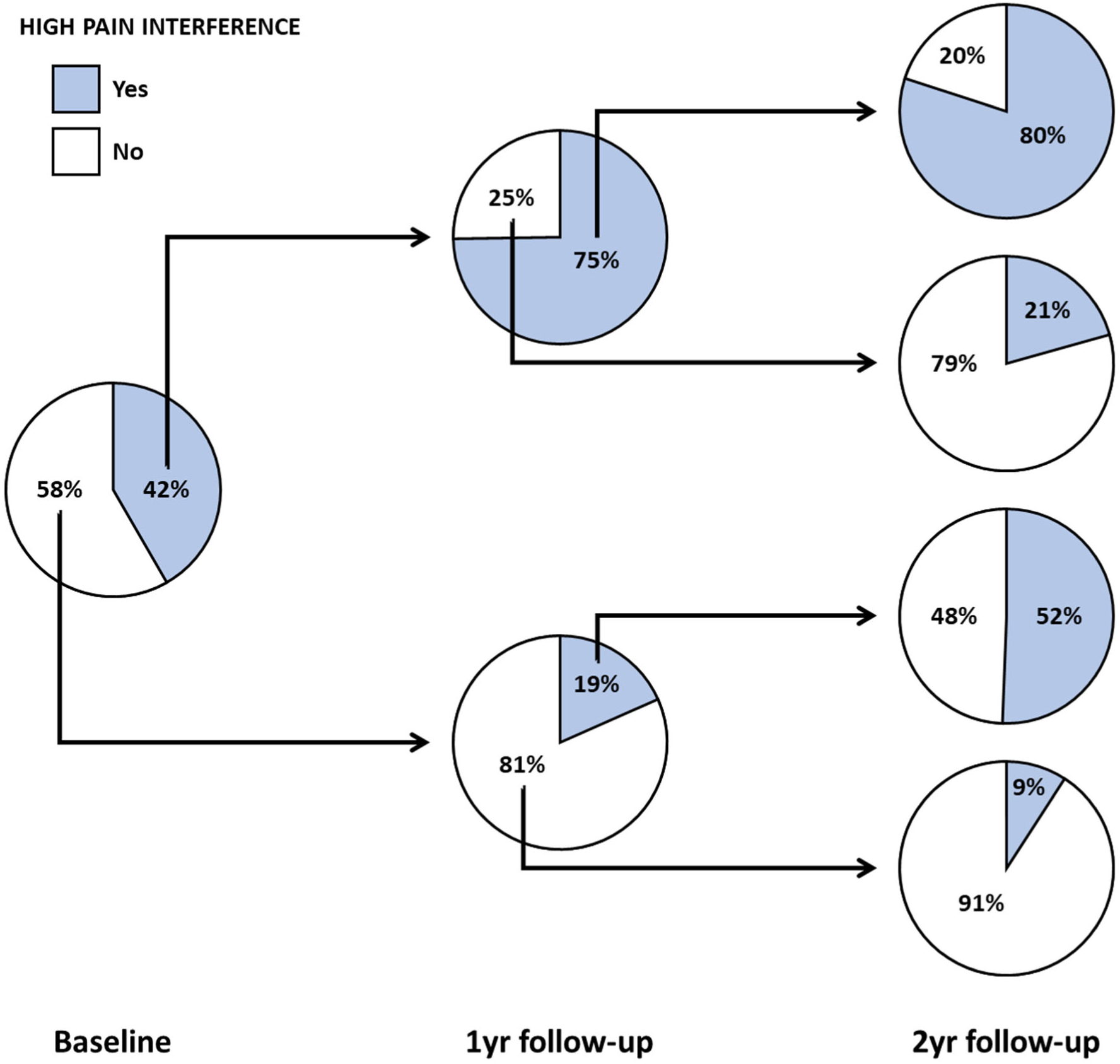
Longitudinal trajectory high pain interference.

**Supplementary Figure 3:**
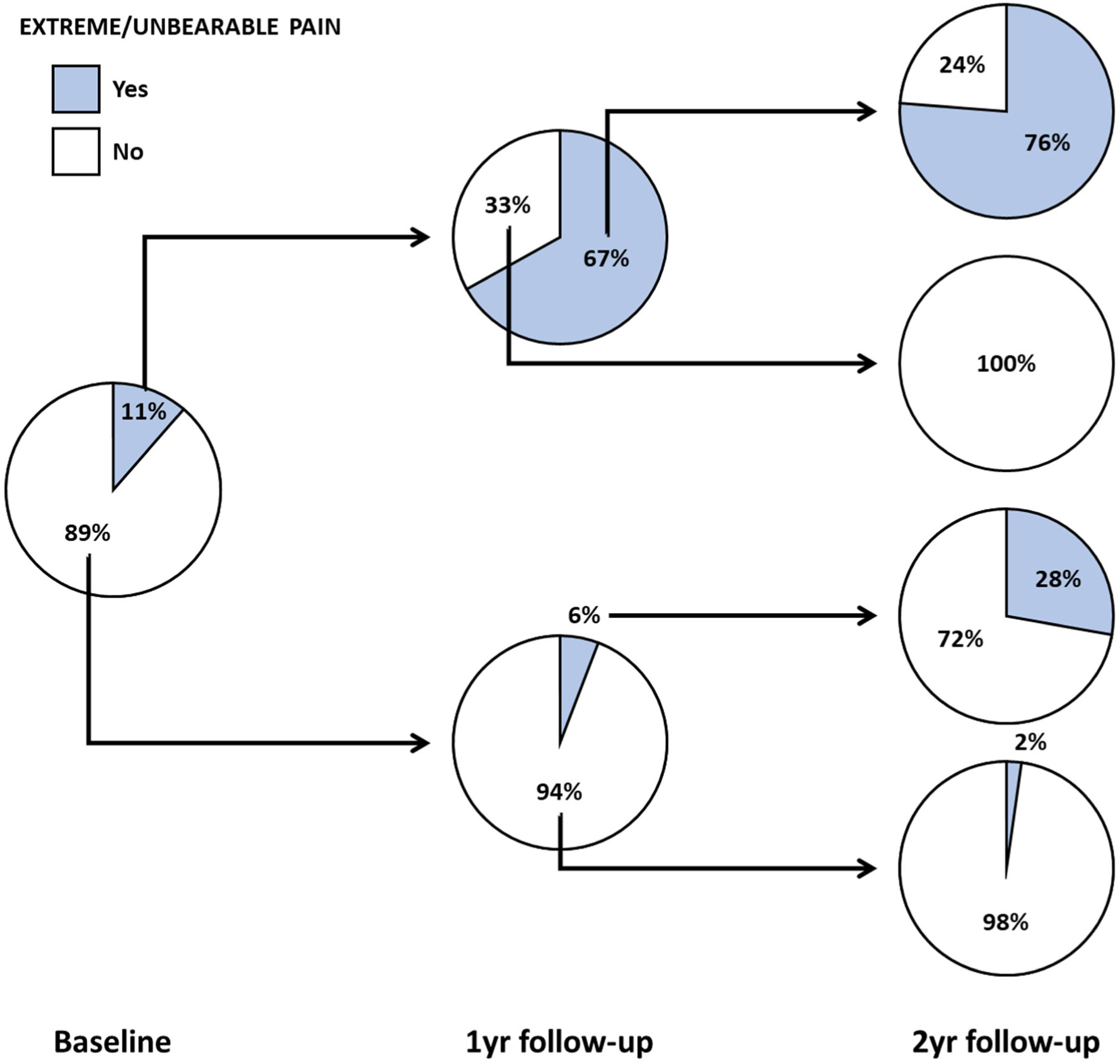
Longitudinal trajectory of extreme/unbearable pain.

**Supplementary Table 1:**
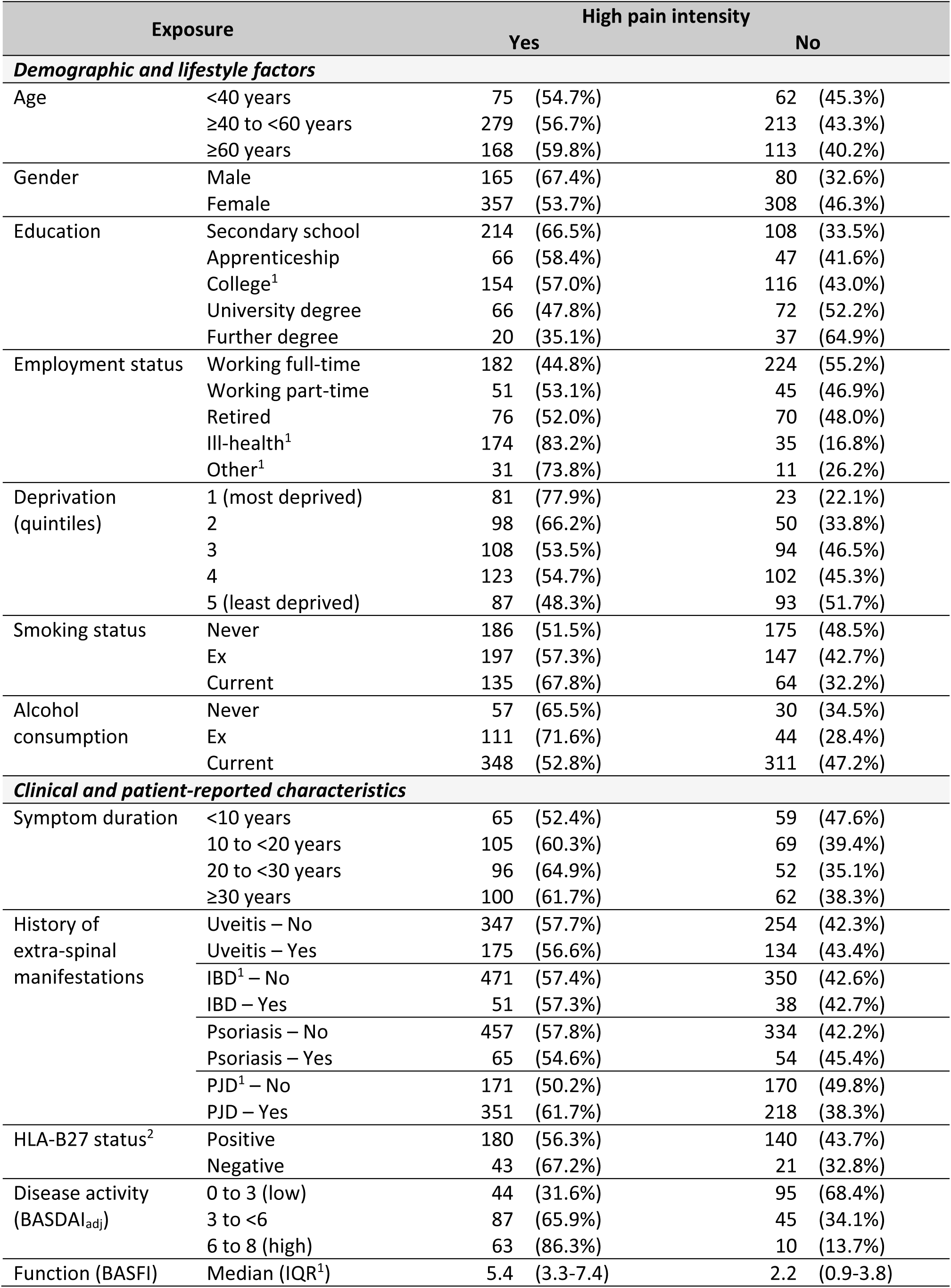
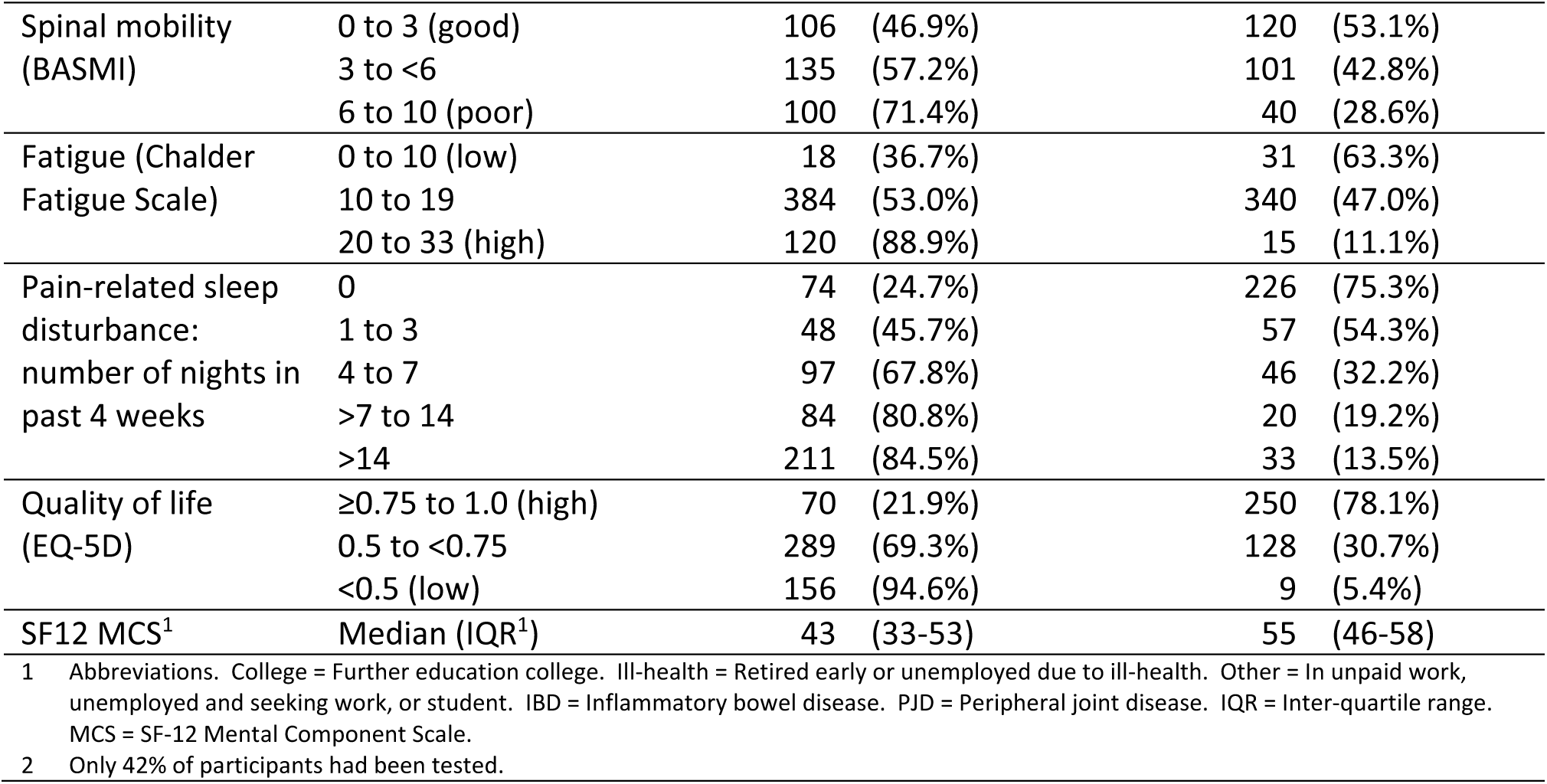
Characteristics of participants – high pain severity.

| Exposure |  | High pain intensity |  |
| --- | --- | --- | --- |
|  |  | Yes | No |
| ***Demographic and lifestyle factors*** |  |  |  |
| Age | <40 years | 75 (54.7%) | 62 (45.3%) |
|  | ≥40 to <60 years | 279 (56.7%) | 213 (43.3%) |
|  | ≥60 years | 168 (59.8%) | 113 (40.2%) |
| Gender | Male | 165 (67.4%) | 80 (32.6%) |
|  | Female | 357 (53.7%) | 308 (46.3%) |
| Education | Secondary school | 214 (66.5%) | 108 (33.5%) |
|  | Apprenticeship | 66 (58.4%) | 47 (41.6%) |
|  | College<sup>1</sup> | 154 (57.0%) | 116 (43.0%) |
|  | University degree | 66 (47.8%) | 72 (52.2%) |
|  | Further degree | 20 (35.1%) | 37 (64.9%) |
| Employment status | Working full-time | 182 (44.8%) | 224 (55.2%) |
|  | Working part-time | 51 (53.1%) | 45 (46.9%) |
|  | Retired | 76 (52.0%) | 70 (48.0%) |
|  | Ill-health<sup>1</sup> | 174 (83.2%) | 35 (16.8%) |
|  | Other<sup>1</sup> | 31 (73.8%) | 11 (26.2%) |
| Deprivation (quintiles) | 1 (most deprived) | 81 (77.9%) | 23 (22.1%) |
|  | 2 | 98 (66.2%) | 50 (33.8%) |
|  | 3 | 108 (53.5%) | 94 (46.5%) |
|  | 4 | 123 (54.7%) | 102 (45.3%) |
|  | 5 (least deprived) | 87 (48.3%) | 93 (51.7%) |
| Smoking status | Never | 186 (51.5%) | 175 (48.5%) |
|  | Ex | 197 (57.3%) | 147 (42.7%) |
|  | Current | 135 (67.8%) | 64 (32.2%) |
| Alcohol consumption | Never | 57 (65.5%) | 30 (34.5%) |
|  | Ex | 111 (71.6%) | 44 (28.4%) |
|  | Current | 348 (52.8%) | 311 (47.2%) |
| ***Clinical and patient-reported characteristics*** |  |  |  |
| Symptom duration | <10 years | 65 (52.4%) | 59 (47.6%) |
|  | 10 to <20 years | 105 (60.3%) | 69 (39.4%) |
|  | 20 to <30 years | 96 (64.9%) | 52 (35.1%) |
|  | ≥30 years | 100 (61.7%) | 62 (38.3%) |
| History of extra-spinal manifestations | Uveitis – No | 347 (57.7%) | 254 (42.3%) |
|  | Uveitis – Yes | 175 (56.6%) | 134 (43.4%) |
|  | IBD<sup>1</sup> – No | 471 (57.4%) | 350 (42.6%) |
|  | IBD – Yes | 51 (57.3%) | 38 (42.7%) |
|  | Psoriasis – No | 457 (57.8%) | 334 (42.2%) |
|  | Psoriasis – Yes | 65 (54.6%) | 54 (45.4%) |
|  | PJD<sup>1</sup> – No | 171 (50.2%) | 170 (49.8%) |
|  | PJD – Yes | 351 (61.7%) | 218 (38.3%) |
| HLA-B27 status<sup>2</sup> | Positive | 180 (56.3%) | 140 (43.7%) |
|  | Negative | 43 (67.2%) | 21 (32.8%) |
| Disease activity (BASDAI<sub>adj</sub>) | 0 to 3 (low) | 44 (31.6%) | 95 (68.4%) |
|  | 3 to <6 | 87 (65.9%) | 45 (34.1%) |
|  | 6 to 8 (high) | 63 (86.3%) | 10 (13.7%) |
| Function (BASFI) | Median (IQR<sup>1</sup>) | 5.4 (3.3-7.4) | 2.2 (0.9-3.8) |

|  |  |  |  |  |  |
| --- | --- | --- | --- | --- | --- |
| Spinal mobility (BASMI) | 0 to 3 (good) | 106 | (46.9%) | 120 | (53.1%) |
|  | 3 to <6 | 135 | (57.2%) | 101 | (42.8%) |
|  | 6 to 10 (poor) | 100 | (71.4%) | 40 | (28.6%) |
| Fatigue (Chalder Fatigue Scale) | 0 to 10 (low) | 18 | (36.7%) | 31 | (63.3%) |
|  | 10 to 19 | 384 | (53.0%) | 340 | (47.0%) |
|  | 20 to 33 (high) | 120 | (88.9%) | 15 | (11.1%) |
| Pain-related sleep disturbance: number of nights in past 4 weeks | 0 | 74 | (24.7%) | 226 | (75.3%) |
|  | 1 to 3 | 48 | (45.7%) | 57 | (54.3%) |
|  | 4 to 7 | 97 | (67.8%) | 46 | (32.2%) |
|  | >7 to 14 | 84 | (80.8%) | 20 | (19.2%) |
|  | >14 | 211 | (84.5%) | 33 | (13.5%) |
| Quality of life (EQ-5D) | ≥0.75 to 1.0 (high) | 70 | (21.9%) | 250 | (78.1%) |
|  | 0.5 to <0.75 | 289 | (69.3%) | 128 | (30.7%) |
|  | <0.5 (low) | 156 | (94.6%) | 9 | (5.4%) |
| SF12 MCS<sup>1</sup> | Median (IQR<sup>1</sup>) | 43 | (33-53) | 55 | (46-58) |

Abbreviations. College = Further education college. Ill-health = Retired early or unemployed due to ill-health. Other = In unpaid work, unemployed and seeking work, or student. IBD = Inflammatory bowel disease. PJD = Peripheral joint disease. IQR = Inter-quartile range. MCS = SF-12 Mental Component Scale.
2 Only 42% of participants had been tested.

**Supplementary Table 2:**
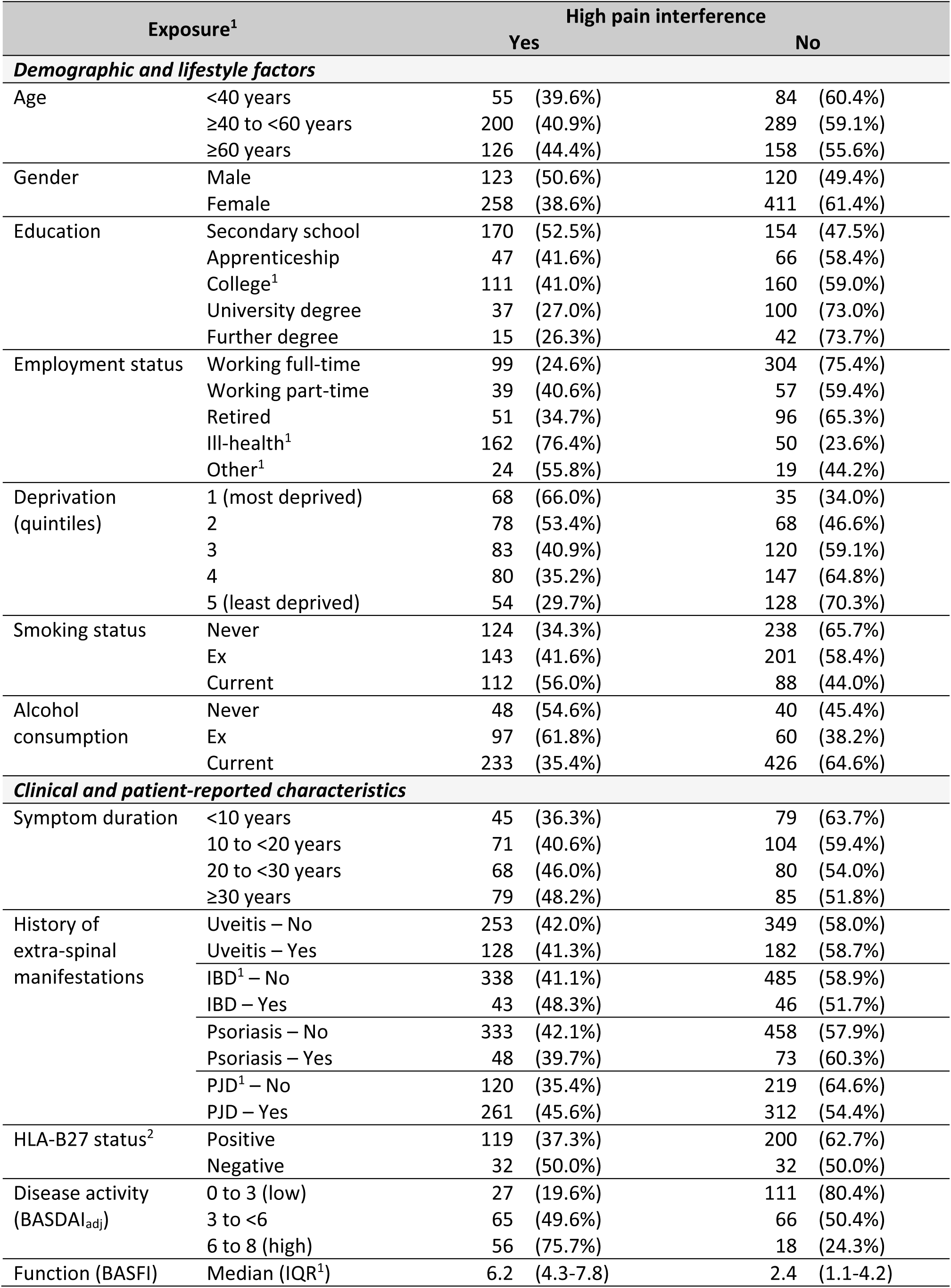
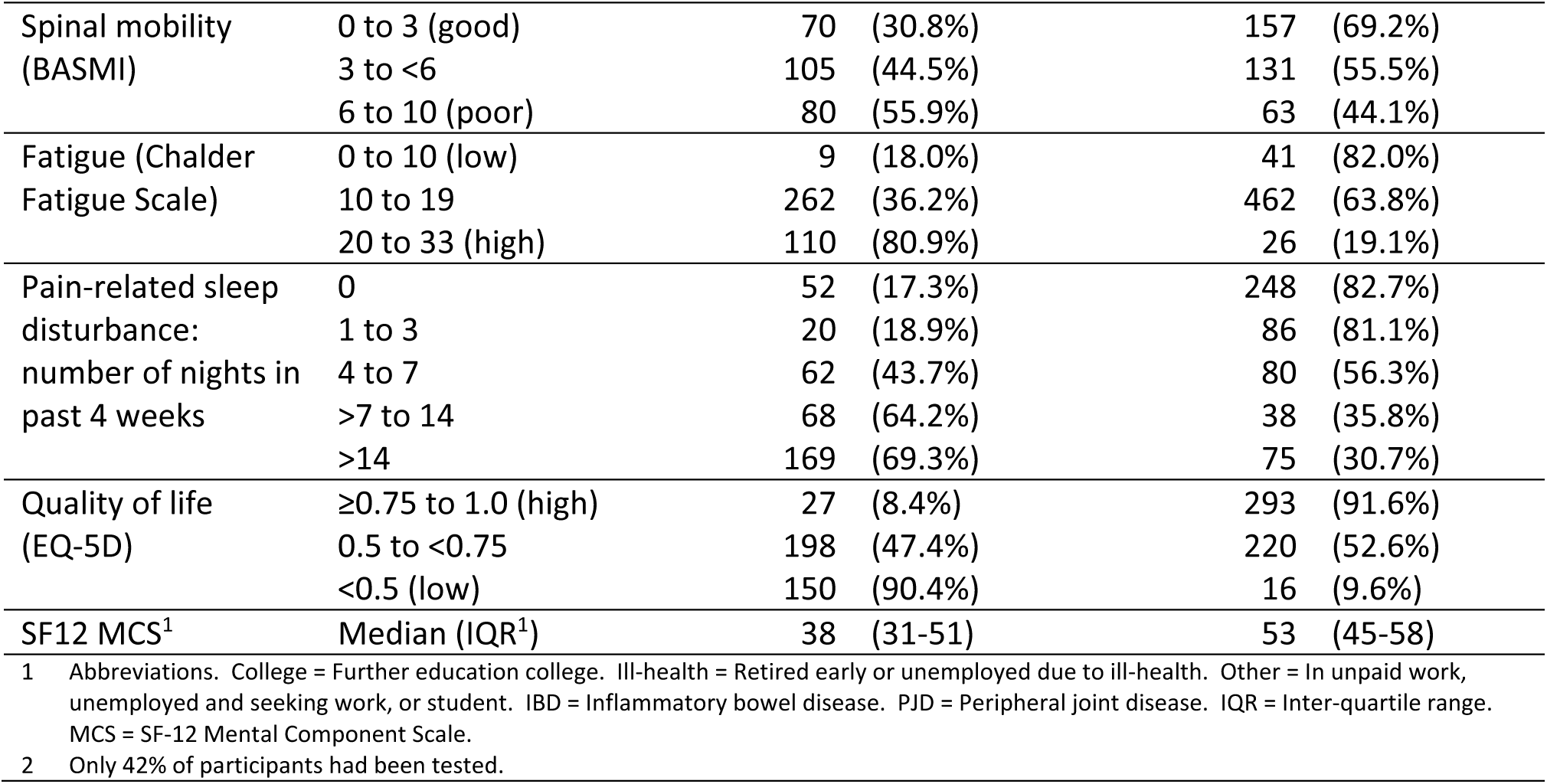
Characteristics of participants – high pain interference.

| Exposure<sup>1</sup> |  | High pain interference |  |
| --- | --- | --- | --- |
|  |  | Yes | No |
| ***Demographic and lifestyle factors*** |  |  |  |
| Age | <40 years | 55 (39.6%) | 84 (60.4%) |
|  | ≥40 to <60 years | 200 (40.9%) | 289 (59.1%) |
|  | ≥60 years | 126 (44.4%) | 158 (55.6%) |
| Gender | Male | 123 (50.6%) | 120 (49.4%) |
|  | Female | 258 (38.6%) | 411 (61.4%) |
| Education | Secondary school | 170 (52.5%) | 154 (47.5%) |
|  | Apprenticeship | 47 (41.6%) | 66 (58.4%) |
|  | College<sup>1</sup> | 111 (41.0%) | 160 (59.0%) |
|  | University degree | 37 (27.0%) | 100 (73.0%) |
|  | Further degree | 15 (26.3%) | 42 (73.7%) |
| Employment status | Working full-time | 99 (24.6%) | 304 (75.4%) |
|  | Working part-time | 39 (40.6%) | 57 (59.4%) |
|  | Retired | 51 (34.7%) | 96 (65.3%) |
|  | Ill-health<sup>1</sup> | 162 (76.4%) | 50 (23.6%) |
|  | Other<sup>1</sup> | 24 (55.8%) | 19 (44.2%) |
| Deprivation (quintiles) | 1 (most deprived) | 68 (66.0%) | 35 (34.0%) |
|  | 2 | 78 (53.4%) | 68 (46.6%) |
|  | 3 | 83 (40.9%) | 120 (59.1%) |
|  | 4 | 80 (35.2%) | 147 (64.8%) |
|  | 5 (least deprived) | 54 (29.7%) | 128 (70.3%) |
| Smoking status | Never | 124 (34.3%) | 238 (65.7%) |
|  | Ex | 143 (41.6%) | 201 (58.4%) |
|  | Current | 112 (56.0%) | 88 (44.0%) |
| Alcohol consumption | Never | 48 (54.6%) | 40 (45.4%) |
|  | Ex | 97 (61.8%) | 60 (38.2%) |
|  | Current | 233 (35.4%) | 426 (64.6%) |
| ***Clinical and patient-reported characteristics*** |  |  |  |
| Symptom duration | <10 years | 45 (36.3%) | 79 (63.7%) |
|  | 10 to <20 years | 71 (40.6%) | 104 (59.4%) |
|  | 20 to <30 years | 68 (46.0%) | 80 (54.0%) |
|  | ≥30 years | 79 (48.2%) | 85 (51.8%) |
| History of extra-spinal manifestations | Uveitis – No | 253 (42.0%) | 349 (58.0%) |
|  | Uveitis – Yes | 128 (41.3%) | 182 (58.7%) |
|  | IBD<sup>1</sup> – No | 338 (41.1%) | 485 (58.9%) |
|  | IBD – Yes | 43 (48.3%) | 46 (51.7%) |
|  | Psoriasis – No | 333 (42.1%) | 458 (57.9%) |
|  | Psoriasis – Yes | 48 (39.7%) | 73 (60.3%) |
|  | PJD<sup>1</sup> – No | 120 (35.4%) | 219 (64.6%) |
|  | PJD – Yes | 261 (45.6%) | 312 (54.4%) |
| HLA-B27 status<sup>2</sup> | Positive | 119 (37.3%) | 200 (62.7%) |
|  | Negative | 32 (50.0%) | 32 (50.0%) |
| Disease activity (BASDAI<sub>adj</sub>) | 0 to 3 (low) | 27 (19.6%) | 111 (80.4%) |
|  | 3 to <6 | 65 (49.6%) | 66 (50.4%) |
|  | 6 to 8 (high) | 56 (75.7%) | 18 (24.3%) |
| Function (BASFI) | Median (IQR<sup>1</sup>) | 6.2 (4.3-7.8) | 2.4 (1.1-4.2) |

|  |  |  |  |  |  |
| --- | --- | --- | --- | --- | --- |
| Spinal mobility (BASMI) | 0 to 3 (good) | 70 | (30.8%) | 157 | (69.2%) |
|  | 3 to <6 | 105 | (44.5%) | 131 | (55.5%) |
|  | 6 to 10 (poor) | 80 | (55.9%) | 63 | (44.1%) |
| Fatigue (Chalder Fatigue Scale) | 0 to 10 (low) | 9 | (18.0%) | 41 | (82.0%) |
|  | 10 to 19 | 262 | (36.2%) | 462 | (63.8%) |
|  | 20 to 33 (high) | 110 | (80.9%) | 26 | (19.1%) |
| Pain-related sleep disturbance: number of nights in past 4 weeks | 0 | 52 | (17.3%) | 248 | (82.7%) |
|  | 1 to 3 | 20 | (18.9%) | 86 | (81.1%) |
|  | 4 to 7 | 62 | (43.7%) | 80 | (56.3%) |
|  | >7 to 14 | 68 | (64.2%) | 38 | (35.8%) |
|  | >14 | 169 | (69.3%) | 75 | (30.7%) |
| Quality of life (EQ-5D) | ≥0.75 to 1.0 (high) | 27 | (8.4%) | 293 | (91.6%) |
|  | 0.5 to <0.75 | 198 | (47.4%) | 220 | (52.6%) |
|  | <0.5 (low) | 150 | (90.4%) | 16 | (9.6%) |
| SF12 MCS<sup>1</sup> | Median (IQR<sup>1</sup>) | 38 | (31-51) | 53 | (45-58) |

Abbreviations. College = Further education college. Ill-health = Retired early or unemployed due to ill-health. Other = In unpaid work, unemployed and seeking work, or student. IBD = Inflammatory bowel disease. PJD = Peripheral joint disease. IQR = Inter-quartile range. MCS = SF-12 Mental Component Scale.
2 Only 42% of participants had been tested.

**Supplementary Table 3:** Characteristics of participants – extreme / unbearable pain.

| Exposure<sup>1</sup> |  | High pain intensity |  |
| --- | --- | --- | --- |
|  |  | Yes | No |
| ***Demographic and lifestyle factors*** |  |  |  |
| Age | <40 years | 19 (13.4%) | 123 (86.6%) |
|  | ≥40 to <60 years | 63 (12.8%) | 429 (87.2%) |
|  | ≥60 years | 23 (8.2%) | 259 (91.8%) |
| Gender | Male | 38 (15.8%) | 203 (84.2%) |
|  | Female | 67 (9.9%) | 608 (90.1%) |
| Education | Secondary school | 59 (18.2%) | 266 (81.8%) |
|  | Apprenticeship | 14 (12.5%) | 98 (87.5%) |
|  | College<sup>1</sup> | 23 (8.4%) | 250 (91.6%) |
|  | University degree | 6 (4.3%) | 133 (95.7%) |
|  | Further degree | 3 (5.3%) | 54 (94.7%) |
| Employment status | Working full-time | 18 (4.4%) | 390 (95.6%) |
|  | Working part-time | 13 (13.5%) | 83 (86.5%) |
|  | Retired | 12 (8.0%) | 138 (92.0%) |
|  | Ill-health<sup>1</sup> | 56 (26.9%) | 152 (73.1%) |
|  | Other<sup>1</sup> | 4 (9.1%) | 40 (90.9%) |
| Deprivation (quintiles) | 1 (most deprived) | 18 (17.7%) | 84 (82.3%) |
|  | 2 | 25 (17.4%) | 119 (82.6%) |
|  | 3 | 19 (9.2%) | 188 (90.8%) |
|  | 4 | 27 (11.8%) | 201 (88.2%) |
|  | 5 (least deprived) | 13 (7.0%) | 172 (93.0%) |
| Smoking status | Never | 30 (8.2%) | 335 (91.8%) |
|  | Ex | 35 (10.2%) | 309 (89.8%) |
|  | Current | 40 (19.9%) | 161 (80.1%) |
| Alcohol consumption | Never | 18 (20.4%) | 70 (79.6%) |
|  | Ex | 35 (22.7%) | 119 (77.3%) |
|  | Current | 50 (7.5%) | 614 (92.5%) |
| ***Clinical and patient-reported characteristics*** |  |  |  |
| Symptom duration | <10 years | 7 (5.6%) | 118 (94.4%) |
|  | 10 to <20 years | 18 (10.2%) | 158 (89.8%) |
|  | 20 to <30 years | 16 (10.8%) | 132 (89.2%) |
|  | ≥30 years | 19 (11.5%) | 146 (88.5%) |
| History of extra-spinal manifestations | Uveitis – No | 75 (12.5%) | 525 (87.5%) |
|  | Uveitis – Yes | 30 (9.5%) | 286 (90.5%) |
|  | IBD<sup>1</sup> – No | 94 (11.3%) | 735 (88.7%) |
|  | IBD – Yes | 11 (12.6%) | 76 (87.4%) |
|  | Psoriasis – No | 97 (12.2%) | 696 (87.8%) |
|  | Psoriasis – Yes | 8 (6.5%) | 115 (93.5%) |
|  | PJD<sup>1</sup> – No | 34 (9.9%) | 308 (90.1%) |
|  | PJD – Yes | 71 (12.4%) | 503 (87.6%) |
| HLA-B27 status<sup>2</sup> | Positive | 29 (9.1%) | 290 (90.9%) |
|  | Negative | 14 (21.9%) | 50 (78.1%) |
| Disease activity (BASDAI<sub>adj</sub>) | 0 to 3 (low) | 6 (4.4%) | 132 (95.6%) |
|  | 3 to <6 | 10 (7.6%) | 122 (92.4%) |
|  | 6 to 8 (high) | 30 (40.0%) | 45 (60.0%) |
| Function (BASFI) | Median (IQR<sup>1</sup>) | 6.8 (4.9-8.3) | 3.4 (1.5-6.0) |
| Spinal mobility (BASMI) | 0 to 3 (good) | 18 (7.9%) | 211 (92.1%) |
|  | 3 to <6 | 33 (13.8%) | 206 (86.2%) |
|  | 6 to 10 (poor) | 19 (13.2%) | 125 (86.8%) |
| Fatigue (Chalder Fatigue Scale) | 0 to 10 (low) | 0 (0%) | 50 (100%) |
|  | 10 to 19 | 51 (7.0%) | 678 (93.0%) |
|  | 20 to 33 (high) | 54 (40.3%) | 80 (59.7%) |
| Pain-related sleep disturbance: number of nights in past 4 weeks | 0 | 2 (0.7%) | 303 (99.3%) |
|  | 1 to 3 | 0 (0%) | 106 (100%) |
|  | 4 to 7 | 8 (5.6%) | 136 (94.4%) |
|  | >7 to 14 | 19 (18.3%) | 85 (81.7%) |
|  | >14 | 75 (31.0%) | 167 (69.0%) |
| Quality of life (EQ-5D) | ≥0.75 to 1.0 (high) | 0 (0%) | 323 (100%) |
|  | 0.5 to <0.75 | 0 (0%) | 427 (100%) |
|  | <0.5 (low) | 104 (65.8%) | 54 (34.2%) |
| SF12 MCS<sup>1</sup> | Median (IQR<sup>1</sup>) | 32 (27-40) | 51 (40-57) |

Abbreviations. College = Further education college. Ill-health = Retired early or unemployed due to ill-health. Other = In unpaid work, unemployed and seeking work, or student. IBD = Inflammatory bowel disease. PJD = Peripheral joint disease. IQR = Inter-quartile range. MCS = SF-12 Mental Component Scale.
2 Only 42% of participants had been tested.

